# Development and validation of radiomics nomograms for preoperative prediction of characteristics in non-small cell lung cancer and circulating tumor cells

**DOI:** 10.1101/2022.02.11.22270861

**Authors:** Yang Wang, Junkai Zhu, Wenxuan Cheng, Li Xu, Xin Wang, Jian Wang, Jun Yang, Fengnan Niu, Wenyi li, Zhibo Wen, Xiaofan Lu, Fangrong Yan

**Author notes:** **Correspondence to:** Dr. Wenyi Li, Ph.D., Department of Endocrinology, Tongren Hospital, Shanghai Jiao Tong University School of Medicine, 1111 XianXia Road, Shanghai 200336, P.R. China., Prof. Zhibo Wen, M.D., Department of Radiology, Zhujiang Hospital, Southern Medical University, 253 Gongye Middle Avenue, Haizhu District, Guangzhou, Guangdong, 510282, P.R. China., Dr. Xiaofan Lu, Ph.D., State Key Laboratory of Natural Medicines, Research Center of Biostatistics and Computational Pharmacy, China Pharmaceutical University, Nanjing 210009, P.R. China., Prof. Fangrong Yan, Ph.D., State Key Laboratory of Natural Medicines, Research Center of Biostatistics and Computational Pharmacy, China Pharmaceutical University, Nanjing 210009, P.R. China. These authors contributed equally (Yang Wang, Junkai Zhu, Wenxuan Cheng). Department of Cancer and Functional Genomics, Institute of Genetics and Molecular and Cellular Biology, CNRS/INSERM/UNISTRA, 67400 Illkirch, France.

## Abstract

**Purpose:** To develop and validate three radiomics nomograms for preoperative prediction of pathological and progression diagnosis in non-small cell lung cancer (NSCLC) as well as circulating tumor cells (CTCs).

**Patients and methods:** A total of 224 and 134 patients diagnosed with NSCLC were respectively gathered in 2018 and 2019 in this study. There were totally 1,197 radiomics features that were extracted and quantified from the images produced by computed tomography (CT). Then we selected the radiomics features with predictive value by LASSO and combined them into radiomics signature. Logistic regression models were built using radiomics signature as the only predictor, which were then converted to nomograms for individualized predictions. Finally, the performance of the nomograms was assessed on both two cohorts.

**Results:** As for discrimination, the AUC of pathological diagnosis nomogram and progression diagnosis nomogram in NSCLC were both higher than 90% in the training cohort and higher than 80% in the validation cohort. The performance of the CTC-diagnosis nomogram was somehow unexpected where the AUC were range from 60% to 70% in both two cohorts. As for calibration, nonsignificant statistics (*p*>0.05) yielded by Hosmer-Lemeshow tests suggested no departure between model prediction and perfect fit. Additionally, decision curve analyses demonstrated the clinically usefulness of the nomograms.

**Conclusion:** We developed radiomics-based nomograms for pathological, progression and CTC diagnosis prediction in NSCLC, respectively. Nomograms for pathological and progression diagnosis were demonstrated well performed to facilitate the preoperative individualized prediction, while the nomogram for CTC-diagnosis prediction needed improvement.

## INTRODUCTION

Lung cancer is the leading cause of cancer-related deaths worldwide, with non-small cell lung cancer (NSCLC) comprising 85% of lung cancers where lung adenocarcinoma (LUAD) is the most common subtype.^1, 2^ Clinical evaluation and staging of NSCLC are used for guiding therapeutic strategy for patients; therefore, accurate diagnosis and staging in the preoperative period is essential for treatment management.^3^ Although several histopathologic findings, such as depth of myometrial invasion, lymphovascular invasion, histologic grade, and nodal status are known to be predictors of NSCLC, these features are only available postoperatively.^4^ Preoperative knowledge of NSCLC can provide valuable information for determining the need for the surgical resection and adjuvant therapy, thus aiding in pretreatment decision making.^5^ Recently, combining circulating tumor cells (CTCs) with other clinical factors was reported to enable reaching high predictive accuracy in the preoperative detection of NSCLC.^6-8^

A circulating tumor cell (CTC) is a cell that has shed into the vasculature or lymphatics from a primary tumor and is carried around the body in the blood circulation. The detection and analysis of CTCs can assist early prognoses evaluation and determine appropriate tailored treatments. For example, the important aspect of the ability to prognose the future disease progression is to, at least temporarily, eliminate the need for a surgery when the repeated CTC counts are low and not increasing. With many advantages, technologies with the requisite sensitivity and reproducibility to detect CTCs in patients with metastatic disease have recently been developed.^28-35^ Although CTC detection is performed via blood tests which are easy and safe to perform and multiple samples can be taken over time, it is challenging with high cost that CTCs are very rare with only a few cells per milliliter of blood and they often express a variety of markers which vary from patient to patient; thus, techniques with high sensitivity and specificity are urgently needed.

Computed tomography (CT) is the first-line imaging method used to evaluate lung cancers in clinical setting;^9^ however, limitations are acknowledged in local staging and in the detection of more subtle cases of pleural invasion.^10, 11^ Recently, radiomics has shown promise in improving models for predicting patient outcomes.^12^ Radiomics is the process of the conversion of medical images into high-dimensional, mineable data via high-throughput extraction of quantitative features, followed by subsequent data analysis for decision support. A central hypothesis demonstrated that radiomics has the potential to enable quantitative measurement of intra- and intertumoral heterogeneity.^13^ Advances in pattern recognition tools and the increase in data set sizes have facilitated the development of radiomics, which may potentially improve predictive accuracy in oncology.^14-17^

Previous studies have shown the combined analysis of a panel of biomarkers, rather than individual analyses, as a signature is the most promising approach that is powerful enough to change clinical management.^19, 20^ Radiomics, which allows the investigation of multiple imaging features in parallel, can provide a combination of features even with biomarkers. Although CT texture assessments by deep learning, have been applied and demonstrated to be useful for prognosis prediction in patients with lung cancer,^21^ an optimal approach that combines multiple laboratory test indicators such as CTCs and imaging biomarkers as a predictive signature has yet to be developed for lung cancer. To the best of our knowledge, there is no evidence determined whether a radiomics signature, or the integration of CTCs and other potentially related clinical features would enable superior prediction of lung cancer as well as whether radiomics signature reflects the CTCs function to a certain extent.

In this context, we therefore aimed to develop and validate radiomics nomograms that incorporated both the radiomics signature and appropriate clinicopathologic risk factors such as CTCs for individualized preoperative prediction of lung cancer on pathological diagnosis (LUAD or not) and disease progression (high tumor grade or not). Additionally, we developed a model to discuss the relationship among CTCs with radiomics signature and other clinical variables. Furthermore, an immunohistochemical correlation analysis was performed between the postoperatively detected expression of specific genes and the dependent variables, which is probably helpful for model validation at genetic level.

## PATIENTS AND METHODS

### Patients

The study enrolled two cohorts which used the same patient recruitment strategy as well as the inclusion and exclusion criteria listed in Data Supplement (Supplementary Materials and Methods). The training cohort contained a total of 224 patients gathered in 2019 with 123 females and 101 males, and the mean age was 58.48 (standard error [SE]: 0.74). The validation cohort incorporated 134 patients gathered in 2018 with 77 females and 57 males, and the mean age was 58.19 (SE: 1.07) (Table 1). Ethical approval was obtained for the retrospective analysis and the level of attrition for the study was consistent with other studies according to the literature.^22^

**Table 1.** Characteristics of patients gathered in this study.

| Parameters | Training Cohort(2019) | Validation Cohort(2018) |
| --- | --- | --- |
| Number of patients | 224 | 134 |
| Age (years) | 58.48 ± 0.74 | 58.19 ± 1.07 |
| Gender (Female : Male) | 1.22:1 (123:101) | 1.35:1 (77:57) |
| CTC (FU) | 10.77 ± 0.28 | 11.91 ± 0.49 |
| CTC result (Positive : Negative) | 2.29:1 (156:68) | 3:1 (99:33) |
| Tumor pathological diagnosis (Lung adenocarcinoma : Lung squamous cell carcinoma : Lung adenosquamous carcinoma : Others) | 211(94.20%):6(2.68%):3(1.34%):4(1.79%) | 124(92.54%):9(6.72%):0(0):1(0.75%) |
| Tumor grade (Low : Low-Medium : Medium : Medium-High : High) | 6(2.82%):20(9.39%):66(30.99%):25(11.74%):96(45.07%) | 7(6.31%):8(7.21%):37(33.33%):15(13.51%):44(39.64%) |
| Short spiculation sign (Positive : Negative) | 2.07:1 (149:72) | 3.03:1 (100:33) |
| Pleural indentation (Positive : Negative) | 1.70:1 (139:82) | 2.24:1 (92:41) |
| Pleural indentation grade (None : Low : Medium : High) | 81(36.65%):96(43.44%):38(17.19%):6(2.71%) | 43(32.33%):75(56.39%):11(8.27%):4(3.01%) |
| Long spiculation sign (Positive : Negative) | 0.91:1 (105:116) | 0.99:1 (66:67) |

\* Continuous values are presented with mean ± standard error of the mean (SEM).
\* Pleural indentation represents the sign of vertical invasion of the tumor into the visceral pleura.
\* Pleural indentation grade is divided according to the number of pleural indentations: None (No pleural indentations), Low (1 or 2 pleural indentations), Medium (3 or 4 pleural indentations), High (5 or more than 5 pleural indentations).

### Dependent variables and dataset partition

Three clinical observation of interest that we would like to fit as dependent variables in this study, including results of LUAD-diagnosis, high tumor grade (HTG) diagnosis and CTC-diagnosis. In this context, three datasets were harnessed separately for model development from totally 358 patients which combined the training and validation cohorts. An independent dataset was also obtained with four tumor invasive variables from 358 patients. To be specific, LUAD-diagnosis came from tumor pathological postoperative diagnosis grouped by LUAD, lung squamous cell carcinoma, lung adenosquamous carcinoma and others (Table 1). Four types of diagnosis results were divided into two categories based on whether the patient had LUAD or not. The dataset was exactly the same as the original dataset with 358 patients where 224 for training cohort and 134 for validation cohort (Supplementary Table S1). Additionally, dataset of HTG-diagnosis was reduced to 324 patients where 213 for training cohort and 111 for validation cohort (Supplementary Table S2) after removing the patients without information of tumor grade. According to the degree of tumor differentiation, tumor grade was divided into 5 ordinal categories: low, low-medium, medium, medium-high and high (Table 1). Patients belonging to medium-high or high category were labelled as HTG, while the remaining were labelled as not HTG. CTC-diagnosis determined by CTC detection value was collected by blood tests on patients with tumors, and the CTC-diagnosis result was defined as positive if the detection value was higher than the threshold of 8.7; otherwise, negative was defined (Table 1). Two patients were removed due to the lack of CTC-diagnosis result and the sample size of such dataset was 356 where 224 for training cohort and 132 for validation cohort (Supplementary Table S3). The independent dataset contained 354 patients where 4 patients were removed due to the lack of images on the four tumor invasive variables.

### Data acquisition and processing

Two kinds of data were used in the study, including image data and clinical detection data; data acquisition and processing were detailed below, respectively. We extracted and quantified a total of 1,197 radiomics features from the image data, and z-score normalization was performed on the training and validation cohorts, respectively. The process of image acquisition, imaging protocol, tumor segmentation along with radiomics feature extraction was described in detail in Data Supplement (Supplementary Materials and Methods). Additionally, for analytic purpose, four tumor invasive features were curated as three indicators and one ordinal categorical variable. Specifically, indicators were signs that whether patients presented with short spiculation sign, pleural indentation or long spiculation sign; one was indicated for specific indicator if positively presented with specific sign and zero was indicated otherwise. The number of pleural indentations was categorized as four levels named pleural indentation grade, including none (no pleural indentations), low (1 or 2 pleural indentations), medium (3 or 4 pleural indentations) and high (5 or more than 5 pleural indentations). Information on four tumor invasive features were collected from the images by radiologists (Figure 1, Table1, Supplementary Table S1-S3). For data collected from clinical setting, value of CTC detection was obtained through blood tests on patients with tumors, and was subsequently converted to CTC-diagnosis result based on the CTC detection value threshold (CTC detection value > 8.7, positive; negative otherwise).

**Figure 1.**
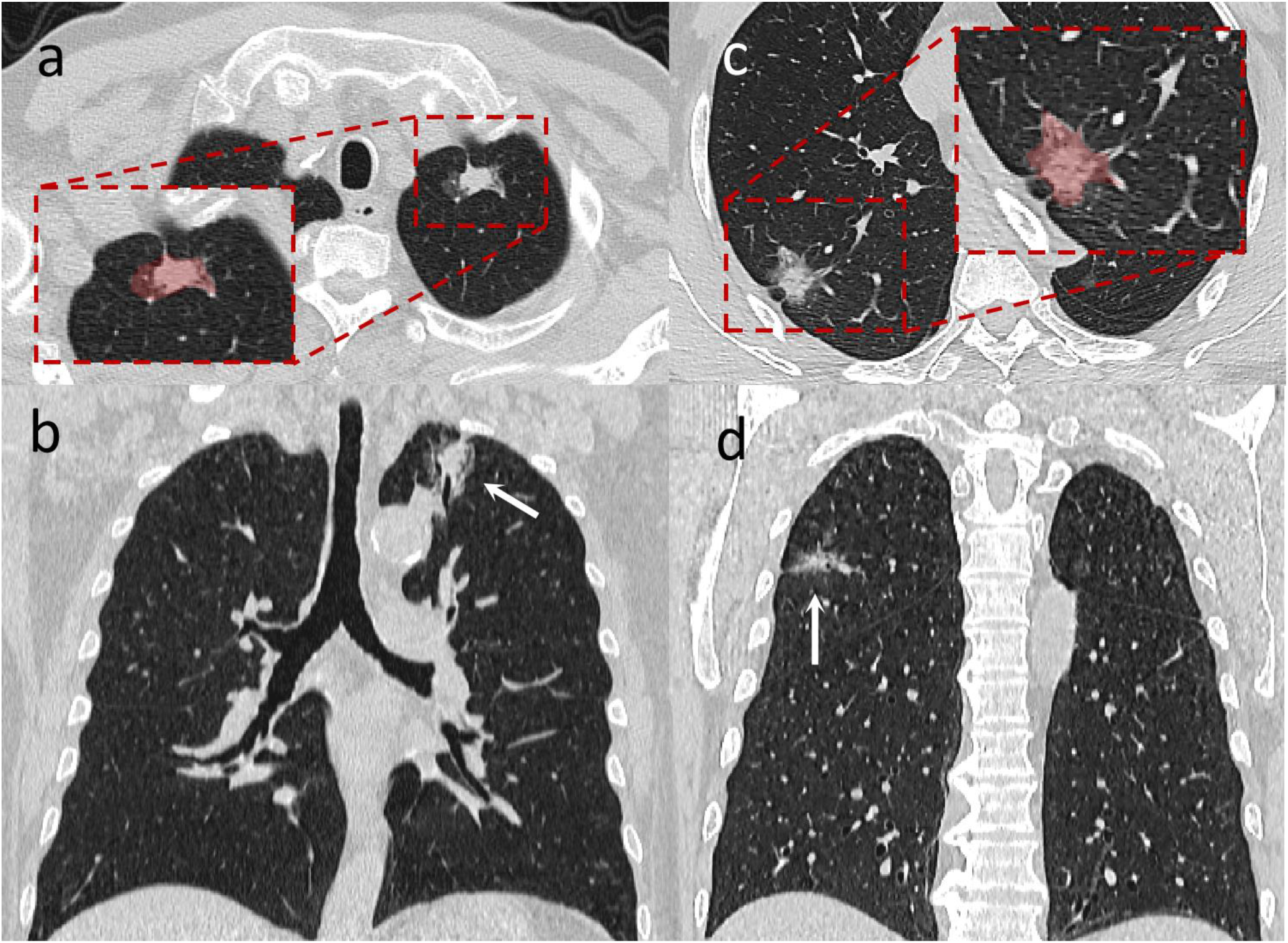
A typical case of lung cancer with a pleural indentation sign and an example of our radiomics outline of a region of interest (ROI) in the left and right lung, respectively. a) The CT images on cross-sectional chest of the left upper lung shows typical signs of pleural indentation and the partial enlarged image shows an example of ROI outline, the light red area. c) In the same patient, the coronal view shows the lesion, and it can be found that the pleural indentation presents signs of invasion at both ends, both ends are nearly 90° to the pleura at the distal end, and the other end points to the pleura in the aortic arch area, and also intersects at an angle of nearly 90°. b) Schematic diagram on the pleural indentation sign of the right lung, it can be seen that there are two tentacles intersecting with the pleura, and the area contained in the ROI outline can be seen by partial enlarged image, the light red area. d) The coronal CT map of the chest in the same patient shows that the pleural indentation sign of the right lung also intersected with the pleura at an angle of nearly 90°.

### Comparison and correlation analysis

Demographic comparisons on continuous preoperative clinical characteristics were performed between different status of dependent variables in two cohorts respectively using Student’s t-test. Correlation analyses (independent test) were conducted among categorical preoperative clinical characteristics with dependent variables in two cohorts respectively by contingency tables (Supplementary Table S1-S3). The relationship between the incidence and cohort of the three dependent variables was further examined.

### Feature selection and radiomics signature building

The least absolute shrinkage and selection operator (LASSO) method^23^ was performed using the R package “glmnet” to select the most predictive features from totally 1,197 radiomics features in the training cohort. Radiomics signature was designed as a vector composed by radiomics score calculated for each patient through a linear combination of selected features that were weighted by their respective coefficients. For example, if there are *p* patients with *q* radiomics features, the radiomics signature is built as the following:

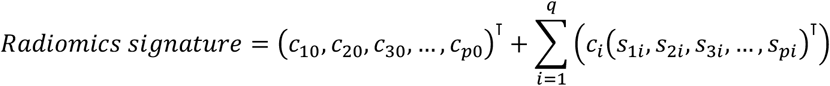

Where *c*_*t* 0_ = *c*_0_ (*t* = 1, 2, 3,…, *p*) and *c*_0_ represents the intercept derived by LASSO method; *c*_*i*_ represents the coefficient of the *i*^th^ radiomics feature derived by LASSO method and *s*_*ti*_ (*t* = 1, 2, 3,…, *p*; *i* = 1, 2, 3…, *q*) represents the value of the *i*^th^ radiomics feature on the *t*^th^ patient.

### Development of predictive models and nomograms on training cohort

Before model development, all the predictor (age, gender, radiomics signature, four tumor invasive variables and CTC-diagnosis result that was just for LUAD as well as HTG diagnosis model development) combinations were tested and compared to select the combination with best performance by Best Subset GLM algorithm (bestglm) using the R package “bestglm” (Supplementary Materials and Methods). To validate the combination selection, model comparisons were performed incorporating different predictor combinations made up of radiomics signature, CTC and four invasive variables to develop models evaluated by the area under curve (AUC) generated from the receiver operating characteristic (ROC) on both two cohorts, respectively. Then, a logistic regression model (Supplementary Materials and Methods) was built between the dependent variable with the selected variables combination. To provide a quantitative tool for individualized prediction, a radiomics nomogram (Supplementary Materials and Methods) was also built on the basis of the developed model using the R package “regplot”.

### Performance evaluation of the nomograms on two cohorts

The logistic regression model developed based on the training cohort was applied to all patients in the validation cohort, with total points for each patient calculated. Using the total points as a factor, logistic regression in the validation cohort was performed. To evaluate the performance of the three developed nomograms, AUC generated from the ROC curve was calculated respectively on both training and validation cohorts using the R package “pROC”. Furthermore, the Hosmer-Lemeshow test was performed using the R package “ResourceSelection”, which implies that the nomogram dose not calibrate perfectly if the test statistic is significant.^24^

### Clinical use

Decision curve analysis was separately conducted using the R package “rmda” to determine the clinical usefulness of the three nomograms by quantifying the net benefits at different threshold probabilities.^25^ To be specific, the net benefit was calculated by subtracting the proportion of all patients who are false positive from the proportion who are true positive, weighting by the relative harm of forgoing treatment compared with the negative consequences of an unnecessary treatment. The relative harm was calculated by 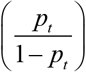 in this condition. *p*_*t*_ (threshold probability) is where the expected benefit of treatment is equal to the expected benefit of avoiding treatment; at which time a patient will opt for treatment informs us of how a patient weighs the relative harms of false-positive results and false-negative results ([*a* − *c*] / [*b* − *d*] = [1− *p*_*t*_] / *p*_*t*_) ; *a* − *c* is the harm from a false-negative result; *b* − *d* is the harm from a false positive result. a, b, c and d give, respectively, the value of true positive, false positive, false negative, and true negative.

### Immunohistochemical correlation analysis

According to the information on the expression of specific genes, immunohistochemical correlation analysis was also performed on three dependent variables as well as the pleural indentation respectively using contingency tables (Supplementary Table S10-S13).

### Statistical analysis

Statistical analysis was conducted with R software (Version 4.0.2). For unadjusted comparisons, a two-sided *p* < 0.05 was considered statistically significant.

## RESULTS

### (1). Model of LUAD-diagnosis

To investigate the relationship among LUAD-diagnosis result with preoperative clinical variables, a model was built for LUAD-diagnosis result prediction.

Firstly, comparison and correlation analyses among preoperative clinical characteristics with LUAD-diagnosis result showed that LUAD-diagnosis result was significantly associated with patients whether they presented sign of long spiculation or not in training cohort (*p*=0.006);in validation cohort we found LUAD-diagnosis result was associated with gender (*p*<0.001), and the age of patients were significantly different (*p*<0.001), which may provide additional value on feature selection and model building (Supplementary Table S1). Besides, there were no significant differences between the two cohorts in view of LUAD incidence (*p*=0.536). LUAD positivity was 94.20% and 92.54% in the training and validation cohorts respectively, indicating a high incidence.

Then, based on the radiomics-related features, 1,197 features were reduced to 9 potential predictors from the training cohort using LASSO (Figure 2a, 2b), which were further harnessed to build a radiomics signature. We observed a significant difference on radiomics score between LUAD-diagnosis results in two cohorts (both, *p*<0.001). Patients diagnosed with LUAD generally had higher radiomics score in two cohorts (Figure 2c, Supplementary Table S1).

**Figure 2.**
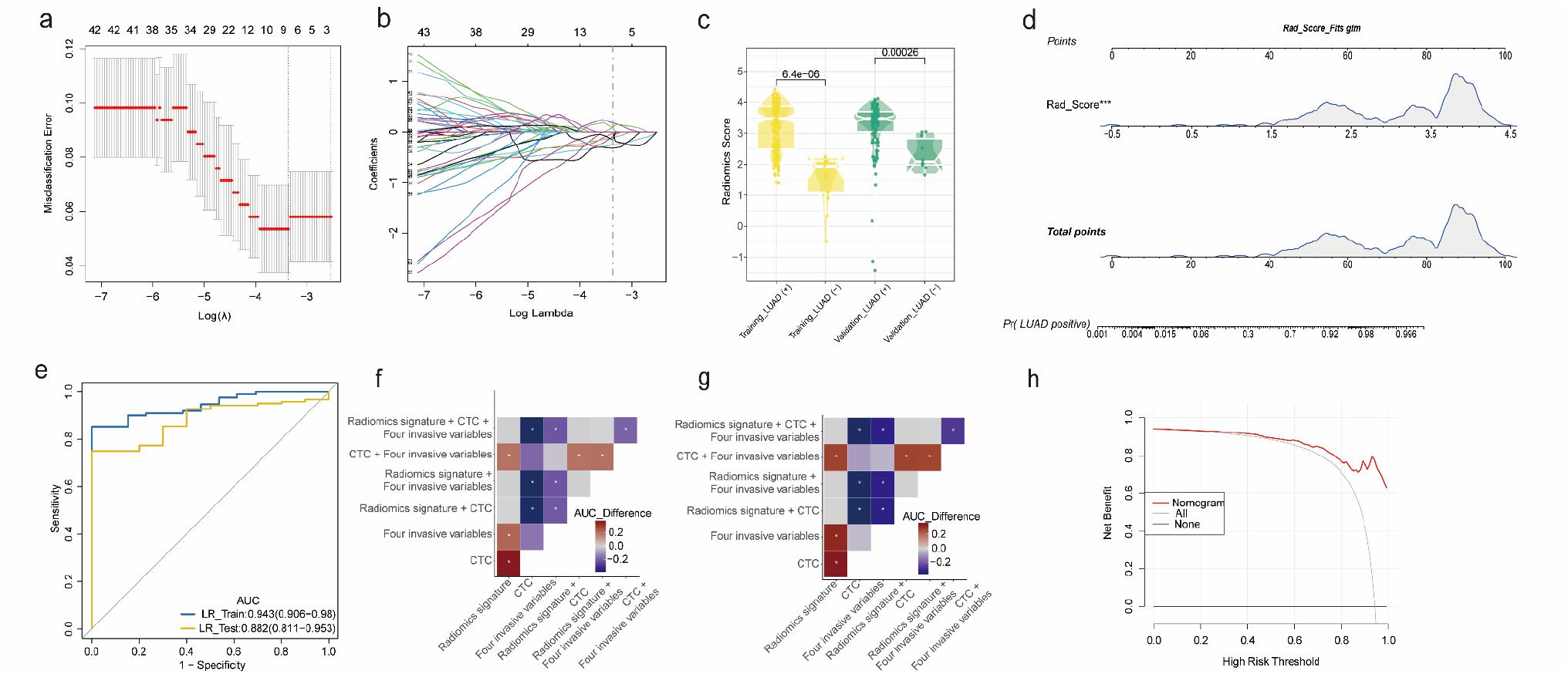
Model of LUAD-diagnosis. a) 10-fold cross-validation via minimum criteria was used to determine the number of the selected radiomics features with nonzero coefficients by LASSO. Misclassification error was plotted versus log(λ). Dotted vertical lines were drawn at the optimal values using the minimum criteria (the left line) and the 1 standard error of the minimum criteria (the 1-SE criteria, the right line). b) A coefficient profile plot was produced against the log(λ) sequence. Vertical lines were drawn at the optimal values using the minimum criteria. Judged by the minimum criteria, a total of 9 features was chosen for LUAD-diagnosis model. c) The boxplot showed a significant difference on radiomics score between different status of LUAD-diagnosis result in both two cohorts (both, *p*<0.001), and patients diagnosed with LUAD generally had higher radiomics score in both two cohorts. d) A radiomics-based nomograms was developed in the training cohort based on the LUAD-diagnosis model using radiomics signature as the only predictor determined by bestglm. e) The performance of the LUAD-diagnosis nomogram was demonstrated good enough when the AUC was 0.943 (95% CI: 0.906-0.980) on training cohort and 0.882 (95% CI: 0.811-0.953) on validation cohort. Furthermore, model comparisons on LUAD-diagnosis result with totally 7 predictor combinations f) in the training cohort as well as g) the validation cohort both showed that the model using radiomics signature as the only predictor performed better than other combinations without radiomics signature, and there was no significant improvement when adding other candidate variables to the radiomics-based model. “AUC_Difference” means the AUC of the model in x-axis subtracts the AUC of the model in y-axis, and the sign of “*” means the difference of AUC is significant (*p*<0.05). h) Decision curve for LUAD-diagnosis nomogram. The x-axis represents the threshold probability and the y-axis measures the net benefit. The red line represents the radiomics nomogram. The gray line represents the assumption that all patients are event positive, while the black line represents the assumption that no patients are event positive. Based on the decision curve, if the threshold probability of a patient or doctor was higher than approximately 20%, it was recommended to use the radiomics nomogram which added more benefit than either the treat-all-patients strategy or the treat-none-patients strategy for prediction.

To develop a predictive model on LUAD-diagnosis result, radiomics signature and other clinical variables were supplied as candidate predictors. Only radiomics signature survived from the bestglm was chosen to develop a logistic regression prediction model (Supplementary Table S4). We then mirrored the model to a nomogram which can be used clinically for individualized prediction (Figure 2d). The results of model comparisons also demonstrated the selection by bestglm, which showed that if the model was incorporated by radiomics signature along with other candidate variables, there was little improvement on model performance assessed by AUC in both two cohorts (Figure 2f, 2g). The details of the model comparisons considering the AUC as well as the corresponding misclassification rate were also displayed in the Data Supplement (Supplementary Table S7).

Furthermore, we evaluated the performance of the model and nomogram on discrimination as well as calibration separately. As for discrimination, the ROC curve demonstrated a good performance on both the two cohorts. The AUC was 0.943 (95% CI: 0.906-0.980) on training cohort and 0.882 (95% CI: 0.811-0.953) on validation cohort (Figure 2e). As for calibration, we yielded a nonsignificant statistic using the Hosmer-Lemeshow test (*p*=0.341, training cohort; *p*=0.573, validation cohort), suggesting a high consistency between model prediction and perfect fit (Table 2).

**Table 2.** *P* values for Hosmer-Lemeshow tests on three diagnosis nomograms.

| Nomograms | Training Cohort | Validation Cohort |
| --- | --- | --- |
| LUAD-diagnosis | 0.341 | 0.573 |
| HTG-diagnosis | 0.662 | 0.696 |
| CTC-diagnosis | 0.789 | 0.085 |

**NOTE.** If $P < 0.05$ , it implies that the nomogram dose not calibrate perfectly.

Considering the clinical usefulness, the decision curve analysis for the model and nomogram demonstrated that if the threshold probability of a patient or doctor was higher than approximately 20%, using the radiomics nomogram to predict LUAD might add more benefit than both the treat-all-patients and the treat-none strategies (Figure 2h).

### (2). Model of HTG-diagnosis

For the determination of the severity of the tumor, a model was developed to predict the HTG-diagnosis result using proper preoperative clinical variables, with a similar process of LUAD-diagnosis model development.

We did not observe significant differences between training and validation two cohorts in HTG incidence (*p*=0.530). Patients with HTG positivity comprised more than half of the population enrolled in this study (56.81% in training cohort and 53.15% in validation cohort). Comparison and correlation analyses among preoperative clinical characteristics with HTG-diagnosis result (Supplementary Table S2) showed that HTG result was tightly associated with tumor invasive characteristics (short spiculation sign, pleural indentation, pleural indentation grade and long spiculation sign) in training cohort (*p*<0.001), while significantly related to gender (*p*<0.001) as well as tumor invasive characteristics (*p*<0.001) except short spiculation sign in validation cohort. Patients’ ages were significantly different between HTG results in both two cohorts (*p*<0.001, training cohort; *p*=0.002, validation cohort).

For feature selection and radiomics signature building, we chose 9 potential predictors from the training cohort (Figure 3a, 3b), which were used to form a radiomics signature. We found a significant difference on radiomics score between HTG results in two cohorts (both, *p*<0.001; Figure 3c, Supplementary Table S2). Likewise, bestglm selected radiomics signature as the only independent variable, which was validated by model comparisons (Figure 3f, 3g, Supplementary Table S8) to develop a prediction model on HTG-diagnosis result (Supplementary Table S5). A prediction nomogram was subsequently built on the basis of the model (Figure 3d).

**Figure 3.**
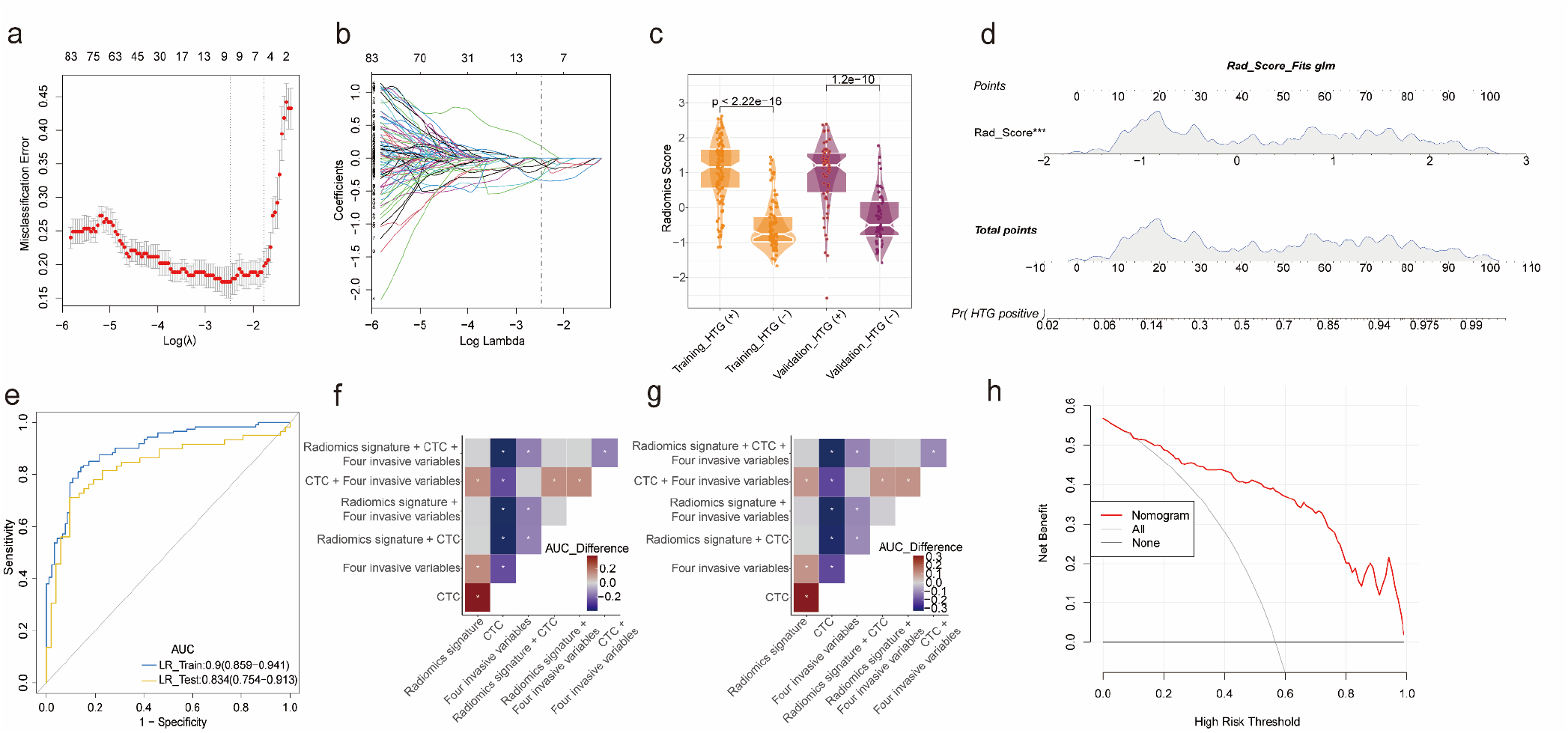
Model of HTG-diagnosis. a). Misclassification error was plotted versus log(λ), and b) a coefficient profile plot was produced against the log(λ) sequence. We selected totally 9 features based on the minimum criteria to develop HTG-diagnosis model. c). Likewise, we also observed a significant difference on radiomics score between HTG results in both two cohorts (both, *p*<0.001) and patients diagnosed with HTG generally had higher radiomics score in both two cohorts. d). We used radiomics signature as the only predictor to develop HTG-diagnosis model in the training cohort and then mirrored the model to a nomogram. e). The AUC was 0.9 (95% CI: 0.859-0.941) on training cohort and 0.834 (95% CI: 0.754-0.913) on validation cohort, which indicated a good performance on both two cohorts. Similarly, in both f) training cohort and g) validation cohort, there were a total of 7 predictor combinations on HTG-diagnosis model comparisons, suggesting a better performance on the model with radiomics signature as the only predictor than other combinations without radiomics signature. However, we found the performance did not improve significantly when other variables joined in the radiomics-based model. h). Decision curve for HTG-diagnosis nomogram, which demonstrated that if the threshold probability of a patient or doctor was higher than roughly 12%, using the radiomics nomogram to predict HTG might add more benefit than both the treat-all-patients and the treat-none strategies.

As for the performance of the model and nomogram, the AUC was 0.9 (95% CI: 0.859-0.941) on training cohort and 0.834 (95% CI: 0.754-0.913) on validation cohort, which indicated a good performance on both two cohorts (Figure 3e). Moreover, the Hosmer-Lemeshow test produced a nonsignificant statistic (*p*=0.662, training cohort; *p*=0.696, validation cohort), suggesting no departure between model prediction and perfect fit (Table 2).

Decision curve analysis demonstrated that if the threshold probability of a patient or doctor was higher than roughly 12%, it was more desirable to use the radiomics nomogram which added more benefit than both the treat-all-patients and the treat-none strategies to predict HTG (Figure 3h).

### (3). Model of CTC-diagnosis

To further investigate the relationship among CTC-diagnosis result with other preoperative clinical variables, we also developed a model on CTC-diagnosis result with proper preoperative clinical variables using the same procedure as the LUAD-diagnosis and HTG-diagnosis model development.

According to the comparison and correlation analyses among preoperative clinical characteristics with CTC-diagnosis result showed that there were no characteristics which were significantly related to CTC result in both two cohorts (Supplementary Table S3). In addition, no statistical differences regarding CTC positive prevalence (*p*=0.279) between training and validation cohorts were observed. CTC positive rate was approximately 0.7 in the population with 69.64% and 75% in the training and validation cohorts, respectively.

Moreover, we selected only 4 potential predictors from the training cohort (Figure 4a, 4b) and combined them into a radiomics signature. A significant difference on radiomics score existed between CTC results just in the training cohort (*p*=0.002), while it was not significantly different in the validation cohort (*p*=0.374). Additionally, patients diagnosed with CTC positive result generally had higher radiomics score in both two cohorts (Figure 4c, Supplementary Table S3).

**Figure 4.**
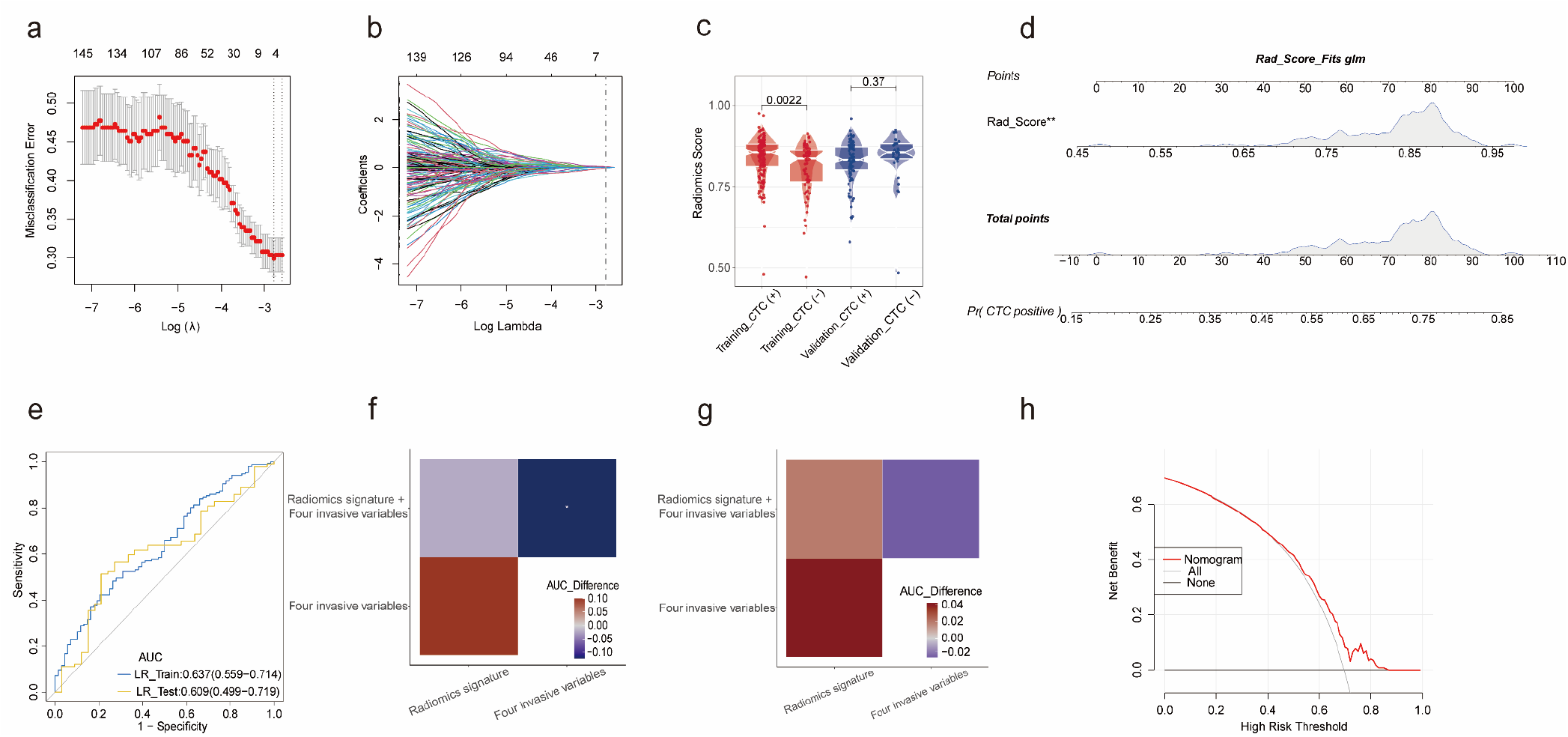
Model of CTC-diagnosis. a). Misclassification error versus log(λ). b) A coefficient profile plot was produced against the log(λ) sequence. We selected only 4 potential features via the minimum criteria and combined them into a radiomics signature for CTC-diagnosis model development. c). A significant difference on radiomics score existed between CTC results just in the training cohort (*p*=0.002), while it was not significantly different in the validation cohort (*p*=0.374). Additionally, patients diagnosed with CTC generally had higher radiomics score in both two cohorts. d). A nomogram was built on the basis of the CTC-diagnosis model, with radiomics signature as the only predictor. e). The performance of the nomogram was worse than the LUAD-diagnosis nomogram as well as the HTG-diagnosis nomogram, which needed improvement. The AUC was 0.637 (95% CI: 0.559-0.714) on training cohort and 0.609 (95% CI: 0.499-0.719) on validation cohort. Moreover, according to the model comparisons with only 3 predictor combinations in f) the training cohort as well as g) the validation cohort, we observed that the model with radiomics signature as the only predictor was slightly better than those without radiomics signature, but the improvement was not significant after mixing four invasive variables into the radiomics-based model. h). Decision curve for CTC-diagnosis nomogram indicated that it was more appropriate to use the radiomics nomogram which added more benefit than both the treat-all-patients and the treat-none strategies to predict CTC-diagnosis result when the threshold probability of a patient or doctor was higher than about 42%.

We then applied bestglm using Bayesian information criterion to determine radiomics signature as the only predictor for model development on CTC-diagnosis prediction (Supplementary Table S6). To confirm the predictor selection, we also performed model comparisons (Figure 4f, 4g, Supplementary Table S9). Simultaneously, a prediction nomogram was also built based on the model (Figure 4d). Unexpectedly, model and nomogram performed worse than the LUAD-diagnosis model as well as the HTG-diagnosis model. The AUC was 0.637 (95% CI: 0.559-0.714) on training cohort and 0.609 (95% CI: 0.499-0.719) on validation cohort (Figure 4e). Additionally, a nonsignificant statistic (*p*=0.789, training cohort; *p*=0.085, validation cohort) yielded by the Hosmer-Lemeshow test suggested no departure between model prediction and perfect fit (Table 2).

For clinical use, it was more appropriate to use the radiomics nomogram which added more benefit than both the treat-all-patients and the treat-none strategies to predict CTC-diagnosis result when the threshold probability of a patient or doctor was higher than about 42% based on the decision curve analysis for the model and nomogram. (Figure 4h).

### (4). Immunohistochemical correlation analysis on research variables

To enhance our findings in genetic level, we performed correlation analysis using immunohistochemical data on LUAD-diagnosis result, which showed that whether gene *TTF-1, P40* or *P63* expressed was significantly related to the incidence of LUAD-diagnosis with a significant statistic separately (*p*<0.001, *TTF-1*; *p*<0.001, *P40*; *p*=0.013, *P63*). As for HTG-diagnosis result, it showed that the expression degree of gene *KI67* was significantly relevant to the tumor differentiation degree which reflects tumor progression, while other genes selected for analysis were of no significant differences (*p*>0.05). At the same time, an analysis was also performed among selected genes with CTC-diagnosis result, which indicated no significant differences as well (*p*>0.05), while analysis on pleural indentation showed that the expression of gene *KI67* (*p*=0.018) and *EGFR* (*p*=0.005) were both significantly associated with the occurrence of pleural indentation (Supplementary Table S10-S13).

## DISCUSSION

Considering the diagnosis of tumor subtype as well as progression, a model along with a nomogram have been developed for LUAD and HTG result prediction respectively, which were both demonstrated to perform well on training and validation cohorts. With the help of the two nomograms, physicians or even patients could be more efficient to make a preoperative individualized prediction to determine whether LUAD occurs and the severity of the disease at the same time, which is in line with the current trend toward personalized medicine.^26^

Nomograms were all based on the radiomics signatures which were built by combining the radiomics features selected by LASSO to calculate the radiomics score. On the one hand, data on radiomics features which comes from images produced by image detection equipment such as CT, magnetic resonance imaging (MRI) and so on can be easily obtained and handled with low cost. On the other hand, as a noninvasive biomarker, radiomics signature is patient-friendly to provide adequate information of great value.

Bestglm was used to select predictors in the process of model development, which tested the performance of all variable combinations and finally chose radiomics signature as the only predictor. Surprisingly, both two models demonstrated adequate discrimination in the two cohorts. The generated AUC for LUAD-diagnosis model was 0.943 and 0.882 on the training and validation cohort respectively with the misclassification rate of 0.049 and 0.105. Likewise, the generated AUC for HTG-diagnosis model was 0.9 and 0.834 on the training and validation cohort separately with the misclassification rate of 0.160 and 0.225. The results of model comparisons also showed that if other current clinical features participated in the two models, the performance would not be better than before. Although the current two models might provide more efficient predictive performance in clinical setting, it is promising for the physicians and researchers to add more clinical characteristics with predictive value to the models and nomograms in practical application.

As a new detection technology that has been popular in recent years, CTC detection has played an important role in providing information on metastatic risk, disease progression and treatment effectiveness, which can help physicians judge the necessity of surgery. Although the CTC detection is noninvasive with great patient compliance, it is challenging with high cost due to the lack of CTCs in blood as well as the variety of markers expressed by CTCs. For this reason, the relationship among CTC-diagnosis result with radiomics signature and other clinical characteristics has also been explored to find out whether CTC detection can be replaced by other detections with low cost. After bestglm variables selection, a model for CTC result prediction was built using radiomics signature as the only predictor. However, the performance of the model was worse than the two models before on both two cohorts. The generated AUC for CTC-diagnosis model was 0.637 and 0.609 on the training and validation cohort respectively with the misclassification rate of 0.281 and 0.280. Furthermore, more potentially relevant clinical variables were incorporated into the model with little performance improved by model comparisons, which may suggest that the CTC detection could be potentially valuable and irreplaceable.

We acknowledged several possible reasons for the unexpected performance of the CTC result diagnosis model. Firstly, confusion matrices showed high false positive rate for predicted results on both two cohorts, which might result from the relatively small sample size. Additionally, the ratio of CTC positive to negative result was 2.29 and 3 in the training and validation cohort respectively; such unbalanced proportions between CTC positive and negative result in two cohorts might cause poor performance as well. Last but not least, it is likely that there was no relationship or the relationship was weak among the CTC result with the current predictors. Therefore, it may be helpful to adopt new potentially relevant predictors to the model instead.

To assess whether the nomogram-assisted decisions would improve patient outcomes than either the treat-all-patients or the treat-none strategy, we applied decision curve analysis in this study instead of multi-institutional prospective validation which may be largely impractical because of the heterogeneity in medical image acquisition and clinical data collection in different institutions. Decision curve analysis focuses on the clinical consequence based on the threshold probability, from which the net benefit could be derived (Net benefit is defined as the proportion of true positives minus the proportion of false positives, weighted by the relative harm of false-positive and false-negative results.).^26, 27^ Decision curves for LUAD, HTG as well as CTC result diagnosis models showed that when the threshold probability of a patient or doctor is higher than 20%, 12% and 42% separately, it is of more benefit to use the nomogram for prediction than both the treat-all-patients and the treat-none schemes.

Interestingly, correlation analyses from genetic level using immunohistochemical data showed that patients with *TTF-1* (+) were demonstrated more likely to be diagnosed with LUAD and patients with *KI67 (+)* or *EGFR (+)* tended to present signs of pleural indentation. In addition, according to the distribution of the four tumor invasive variables in the dataset, it showed that patients who presented signs of short spiculation (70.34%) or pleural indentation (65.25%) were more than those who did not present.

Study limitations included the fact that the sample size was a little small that might not cover enough situations for model training and validation. Besides, the unbalanced proportions of the binary dependent variables in two cohorts such as LUAD-diagnosis result and CTC-diagnosis result might affect the performance of the model training and validation in a way. According to the limitation on data, adjustments are needed for further researches. Moreover, it is feasible to add new potentially related predictors to the models or replace the current ones, which may improve the performance of the models significantly.

To summary, this study introduced three developed radiomics signature-based nomograms for LUAD, HTG and CTC diagnosis result prediction respectively. Nomograms for LUAD and HTG diagnosis result predictions were demonstrated well performed to facilitate the preoperative individualized prediction, while the nomogram for CTC-diagnosis result prediction needed improvement. Additionally, postoperative detection on the expression of specific genes related to the dependent variables were probably useful for validating preoperative model predictions at genetic level.

## Data Availability

All data produced in the present study are available upon reasonable request to the authors.

## SUPPORTING INFORMATION

**Table S1**. Comparison and correlation analyses on LUAD-diagnosis result (+/-) in the training and validation cohorts.

**Table S2**. Comparison and correlation analyses on HTG-diagnosis result (+/-) in the training and validation cohorts.

**Table S3**. Comparison and correlation analyses on CTC-diagnosis result (+/-) in the training and validation cohorts.

**Table S4**. The summary of LUAD-diagnosis model.

**Table S5**. The summary of HTG-diagnosis model.

**Table S6**. The summary of CTC-diagnosis model.

**Table S7**. Details of model comparisons on LUAD-diagnosis result.

**Table S8**. Details of model comparisons on HTG-diagnosis result.

**Table S9**. Details of model comparisons on CTC-diagnosis result.

**Table S10**. Immunohistochemical correlation analysis among LUAD-diagnosis result (+/-) with specific genes.

**Table S11**. Immunohistochemical correlation analysis among HTG-diagnosis result (+/-) with specific genes.

**Table S12**. Immunohistochemical correlation analysis among CTC-diagnosis result (+/-) with specific genes.

**Table S13**. Immunohistochemical correlation analysis among pleural indentation (+/-) with specific genes.

